# Predicting Opportunities for Improvement in Trauma Care: A Registry-Based Cohort Study

**DOI:** 10.1101/2023.01.19.23284654

**Authors:** Jonatan Attergrim, Kelvin Szolnoky, Lovisa Strömmer, Olof Brattström, Gunilla Whilke, Martin Jacobsson, Martin Gerdin Wärnberg

**Author notes:** **Corresponding author: Jonatan Attergrim**, Department of Public Global Health, Karolinska Institutet, 171 77 Stockholm. These authors contributed equally to this work (Jonatan Attergrim, Kelvin Szolnoky).

## Abstract

**Importance:** Trauma quality improvement programs relies on peer review of patient cases to identify opportunities for improvement. Current state-of-the-art systems for selecting patient cases for peer review use audit filters that struggle with poor performance.

**Objective:** To develop models predicting opportunities for improvement in trauma care and compare their performance to currently used audit filters.

**Design, Setting and Participants:** This single-center registry-based cohort study used data from the trauma centre at Karolinska University Hospital in Stockholm, Sweden, between 2013 and 2023. Participants were adult trauma patients included in the local trauma registry. The models predicting opportunities for improvement in trauma care were developed using logistic regression and the eXtreme Gradient Boosting learner (XGBoost) with an add-one-year-in expanding window approach. Performance was measured using the integrated calibration index (ICI), area under the receiver operating curve (AUC), true positive rates (TPR) and false positive rates (FPR). We compared the performance of the models to locally used audit filters.

**Main outcome measure:** Opportunities for improvement, defined as preventable events in patient care with adverse outcomes. These opportunities for improvement were identified by the local peer review processes.

**Results:** A total of 8,220 patients were included. The mean (SD) age was 45 (21), 5696 patients (69%) were male, and the mean (SD) injury severity score was 12 (13). Opportunities for improvement were identified in 496 (6%) patients. The logistic regression and XGBoost models were well calibrated with ICIs (95% CI) of 0.032 (0.032-0.032) and 0.033 (0.032-0.033). Compared to the audit filters, both the logistic regression and XGBoost models had higher AUCs (95% CI) of 0.72 (0.717-0.723) and 0.75 (0.747-0.753), TPR (95% CI) of 0.885 (0.881-0.888) and 0.904 (0.901-0.907), and lower FPR (95% CI) of 0.636 (0.635-0.638) and 0.599 (0.598-0.6). The audit filters had an AUC (95% CI) of 0.616 (0.614-0.618), a TPR (95% CI) of 0.903 (0.9-0.906), and a FPR (95% CI) of 0.671 (0.67-0.672).

**Conclusion and Relevance:** Both the logistic regression and XGBoost models outperformed audit filters in predicting opportunities for improvement among adult trauma patients and can potentially be used to improve systems for selecting patient cases for trauma peer review.

**Key point:** **Question:** How does the performance of machine learning models compare to audit filters when screening for opportunities for improvement, preventable events in care with adverse outcomes, among adult trauma patients?

**Findings:** Our registry-based cohort study including 8,220 patients showed that machine learning models outperform audit filters, with improved discrimination and false-positive rates. Compared to audit filters, these models can be configurated to balance sensitivity against overall screening burden.

**Meaning:** Machine learning models have the potential to reduce false positives when screening for opportunities for improvement in the care of adult trauma patients and thereby enhancing trauma quality improvement programs.

## Introduction

Trauma is a leading cause of death and disability worldwide (1,2). Peer review of patient cases, sometimes referred to as performance improvement, is a critical component of trauma quality improvement programmes (3–5). This review ideally involves representatives from all disciplines and professions involved in trauma care to identify opportunities for improvement, which are preventable events in patient care with adverse outcomes (6).

The current state-of-the-art systems for selecting patient cases for peer review uses audit filters, sometimes in combination with individual human screening (7). Audit filters are sentinel events in patient care that are associated with suboptimal care and potentially poor patient outcomes, such as delays in key interventions or unexpected deaths (3,8). When such an event occurs, it should trigger the peer review process. This process is then followed by the implementation of corrective actions (8).

It has long been known that audit filters perform poorly in this context (9). Replacing filters with trauma mortality prediction models has failed (10–12), likely because they were not developed to predict opportunities for improvement. No published research has evaluated prediction models for opportunities for improvement. We therefore aimed to develop models predicting opportunities for improvement in trauma care and compare their performance to currently used audit filters.

## Methods

### Design

We conducted a registry-based cohort study using all trauma patients included in both the Karolinska University Hospital trauma registry and the trauma care quality database between 2013 and 2022. The study was approved by the Swedish Ethical Review Authority (approval numbers 2021-02541 and 2021-03531).

### Study Setting and Population

The trauma center at the Karolinska University Hospital in Solna, Sweden, manages approximately 1500 acute trauma patients each year (13).

The Karolinska University Hospital trauma registry, part of the Swedish Trauma Registry (13), includes all patients admitted to the Karolinska University Hospital with trauma team activation, regardless of injury severity score (ISS), as well as patients admitted without trauma team activation but found to have ISS of more than 9. The registry includes data on vital signs, times, injuries and interventions and demographics according to the European consensus statement, the Utstein template (14). The care quality database includes data relevant to the peer review process, including audit filters, identified opportunities for improvement, and proposed corrective actions.

The peer review process has evolved over time, but since 2017, a specialized nurse reviews the medical records of all trauma patients and flags patients with potential opportunities for improvement using a set of audit filters (Supplement E2 eTable 1) and clinical experience. A second nurse performs a more in-depth review of all flagged patients. Patients with suspected opportunities for improvement are then reviewed at a multidisciplinary conference, where the final decision on the presence of opportunities for improvement is made. All patients who die are reviewed in a separate conference that evaluates the preventability of the death and determines the presence of any opportunities for improvement. Before 2017, the process was less formalized, and a small group of clinicians involved in trauma care identified opportunities for improvement.

### Eligibility Criteria

We included all patients screened for opportunities for improvement from the trauma registry and trauma care quality database between 1 January 2013 and 31 December 2022. Patients younger than 15 years were excluded because their clinical and review pathways differ from those of adults.

### Outcome

The models’ outcome is the presence of any opportunities for improvement, as determined by the peer review process. The identified opportunities for improvement are further grouped into clinical judgment errors, delays in treatment or diagnosis, missed diagnoses, technical errors, preventable deaths or other errors.

### Sample Size Considerations

The relationship between the number of predictors and required sample size for different learners has not been well researched expect for logistic regression (15,16). We used these guidelines to inform the number of predictors that we could include in our models, and we estimated that with a sample size of 3452, which is equivalent to 80% of the available data from 2017–2020, would support 45 parameters, assuming a 6% event rate, a r2 of 0.11 and a target shrinkage of 0.9.

### Predictors

We selected predictors based on current audit filters, standard demographics, previous research and expert opinion (17). The categorical predictors were gender, type of emergency procedure, highest level of care, reprioritization, type of trauma alarm, discharge destination and death within 30 days. The continuous predictors included age, vital signs on arrival, time to CT and intervention, ISS and length of stay. These final set of predictors comprised 17 variables with 45 corresponding parameters. eTable 2 (Supplement E2) shows all 17 predictors.

### Statistical Analysis Methods

The statistical analyses were conducted using R (18). We developed several prediction models around different learners, available in the supplementary material (Supplement E1 and E2), where we include the best-performing model: eXtreme Gradient Boosting (XGBoost), and the more interpretable model: Logistic Regression. XGBoost builds on the principles of gradient boosting, incorporating various algorithmic optimizations, including parallel tree boosting, to efficiently solve a range of machine learning problems such as classification and regression (19).

To evaluate the models, we used an add-one-year-in expanding window approach to best represents how the models would have performed if implemented prospectively. The years 2017-2022 were all used as separate validation hold-out sets in an iterative fashion. In each iteration, all years prior to the current validation sample were used as training data. The training data were then split, and 80% of the data were used for training and 20% for calibration. We estimated 95% confidence intervals (CIs) for all performance metrics through a bootstrap of 1000 resamples for each validation sample.

#### Data preprocessing and imputation

We developed a preprocessor that rescaled continuous predictors using Yeo-Johnson’s power transformation (20) and recorded categorical predictors into dummy variables via one-hot encoding. Predictors with near-zero variances were excluded. Missing continuous predictors were imputed using the mean of the predictor, and a missing indicator feature was created for each. Categorical predictors were imputed by introducing an ‘unknown’ category. If blood pressure or respiratory rate data were missing but corresponding revised trauma score categorical values were available, we imputed the missing data using the mean of all patients in that category. The preprocessor was initially run on the training sample for each split to learn metrics and prevent data leakage. The trained preprocessor model was then applied independently to both the training and validation samples. To balance the training samples, we used the adaptive synthetic algorithm (21), which generates synthetic data, enabling us to upsample the opportunities for improvement outcomes at a balanced 1:1 ratio between outcome classes.

#### Model development

We developed the logistic regression and XGBoost (19) models using the learners as implemented in the Tidymodels framework (22). All model hyperparameters were optimized on the training sample of each split using five-fold cross-validation through iterative Bayesian optimization, encompassing all the parameters provided by the tidymodels framework.

#### Performance measurements

The prediction models and audit filters performance were assessed and compared in terms of calibration, discrimination, as well as true and false positive rates in each validation sample. Calibration was measured using the integrated calibration index (ICI) (23) and discrimination was measured using the area under the receiver operating characteristic curve (AUC). The ICI was not calculated for the audit filters because they cannot estimate a probability of opportunities for improvement.

To determine the class probability cutoff for the two prediction models model, we first configured them using Platt scaling on a 20% holdout sample from the training samples. We then determined the cutoff that produced a 95% true positive rate on this configuration sample and applied it to the holdout validation sample, called “TPR_95%_”. Additionally, we conducted an analysis to establish an “optimal” cutoff threshold by identifying the point on the ROC curve that maximizes the trade-off between sensitivity and specificity, called “balanced configuration”.

#### Predictor importance

We calculated the predictor importance for the prediction models using permutation feature importance (24) on the nonresampled validation samples. The importance of a feature was thus calculated by taking the average AUC performance when shuffling a feature’s data five times and comparing it to the model’s performance on nonshuffled data.

#### Code availability

The code used in this study is publicly available online: https://github.com/noacs-io/predicting-ofi-in-trauma under the MIT License.

## Results

### Participants

Out of the 13879 patients in the registry included between January 2013 and December 2022, 8220 (59.87%) patients had been reviewed regarding the presence of opportunities for improvement, which were identified in 496 (6%) patients. The most common categories of opportunities for improvement where clinical judgment errors (n=176, 35%) followed by inadequate resources (n=111, 22%). Out of the 718 deaths, 42 (6%) where considered possible preventable making up 9% of all opportunities for improvement. Figure 1 details inclusions and exclusions as well as the frequency of each category of opportunities for improvement. eTable 3 (Supplement E2) shows the specific opportunities for improvement. Patients with opportunities for improvement (mean = 49 years, SD = 21) were slightly older than patients without opportunities for improvement (mean = 45 years, SD = 21). The ISS was greater in patients with opportunities for improvement (mean = 19, SD = 11) than in patients without opportunities for improvement (mean = 12, SD = 13), and patients with opportunities for improvement had longer times (mean = 271 minutes, SD = 323) from hospital arrival to the first major intervention than patients without opportunities for improvement (mean = 251 minutes, SD = 353). Treatment frequencies also differed, with the biggest difference being radiological interventions, where patients with opportunities for improvement (n=32, 6%) had more interventions than those without opportunities for improvement (n=69, 1%). Table 1 shows the characteristics of all included patients.

**Figure 1.**
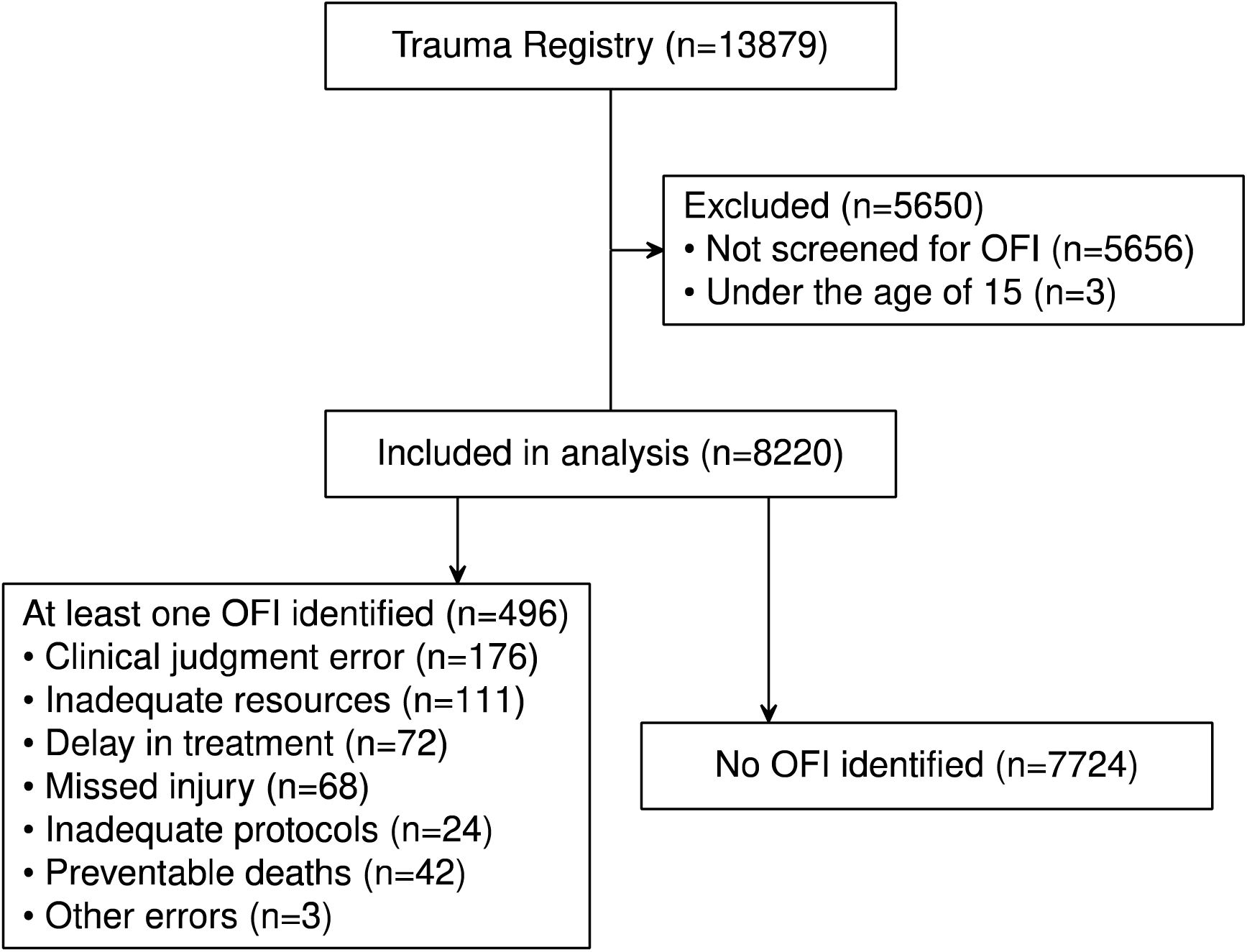
Flowchart describing the exclusions made and the process of accessing trauma patients from arrival until the decision for OFI.

**Table 1.** Demographic and clinical characteristics of patients screened for OFI.

|  | OFI<br>(N=496) | No OFI<br>(N=7724) | Overall<br>(N=8220) |
| --- | --- | --- | --- |
| **Age** |  |  |  |
| Mean (SD) | 49 (22) | 45 (21) | 45 (21) |
| Median [Min, Max] | 49 [15, 97] | 43 [15, 100] | 43 [15, 100] |
| **Sex** |  |  |  |
| Female | 136 (27%) | 2388 (31%) | 2524 (31%) |
| Male | 360 (73%) | 5336 (69%) | 5696 (69%) |
| **Dead at 30 days** |  |  |  |
| Yes | 41 (8%) | 677 (9%) | 718 (9%) |
| No | 453 (91%) | 7038 (91%) | 7491 (91%) |
| Missing | 2 (<1%) | 9 (<1%) | 11 (<1%) |
| **Highest level of care** |  |  |  |
| Emergency department | 22 (4%) | 1467 (19%) | 1489 (18%) |
| General ward | 116 (23%) | 2920 (38%) | 3036 (37%) |
| Operation Theatre | 141 (28%) | 1438 (19%) | 1579 (19%) |
| Specialist ward/Intermediate ward | 50 (10%) | 336 (4%) | 386 (5%) |
| Intensive care unit | 167 (34%) | 1563 (20%) | 1730 (21%) |
| **Injury severity score** |  |  |  |
| Mean (SD) | 19 (11) | 12 (13) | 12 (13) |
| Median [Min, Max] | 17 [0, 75] | 9 [0, 75] | 9 [0, 75] |
| Missing | 0 (0%) | 10 (<1%) | 10 (<1%) |
| **Respiratory rate** |  |  |  |
| Mean (SD) | 19 (5) | 18 (5) | 18 (5) |
| Median [Min, Max] | 18 [0, 40] | 18 [0, 60] | 18 [0, 60] |
| Missing | 51 (10%) | 812 (11%) | 863 (10%) |
| **ED GCS** |  |  |  |
| Mean (SD) | 14 (3) | 14 (2) | 14 (2) |
| Median [Min, Max] | 15 [3, 15] | 15 [3, 15] | 15 [3, 15] |
| Missing | 49 (10%) | 811 (11%) | 860 (10%) |
| **ED Systolic Blood Pressure** |  |  |  |
| Mean (SD) | 133 (34) | 133 (33) | 133 (33) |
| Median [Min, Max] | 135 [0, 237] | 135 [0, 285] | 135 [0, 285] |
| Missing | 0 (0%) | 13 (<1%) | 13 (<1%) |
| **Time to first CT** |  |  |  |
| Mean (SD) | 76 (129) | 71 (159) | 71 (157) |
| Median [Min, Max] | 40 [6, 1339] | 33 [0, 7073] | 33 [0, 7073] |
| Missing | 42 (8%) | 945 (12%) | 987 (12%) |
| **Time to first major intervention** |  |  |  |
| Mean (SD) | 271 (323) | 251 (353) | 253 (349) |
| Median [Min, Max] | 143 [3, 1420] | 102 [0, 2036] | 106 [0, 2036] |
| Missing | 230 (46%) | 5668 (73%) | 5898 (72%) |
| **Emergency procedure** |  |  |  |
| Thoracotomy | 8 (2%) | 97 (1%) | 105 (1%) |
| Laparotomy | 28 (6%) | 213 (3%) | 241 (3%) |
| Pelvis Packing | 0 (0%) | 5 (<1%) | 5 (<1%) |
| Revascularization | 12 (2%) | 37 (<1%) | 49 (1%) |
| Radiological intervention | 32 (6%) | 69 (1%) | 101 (1%) |
| Craniotomy | 42 (8%) | 240 (3%) | 282 (3%) |
| Intracranial pressure measurement | 13 (3%) | 90 (1%) | 103 (1%) |
| Other | 131 (26%) | 1305 (17%) | 1436 (17%) |
| Missing | 230 (46%) | 5668 (73%) | 5898 (72%) |

*Definition of abbreviations:* OFI = Opportunity for Improvement; ED = Emergency Department; GCS = Glasgow Coma Scale. Time to first CT and time to first major intervention: Measured in minutes from arrival at the hospital

**Table 2.**
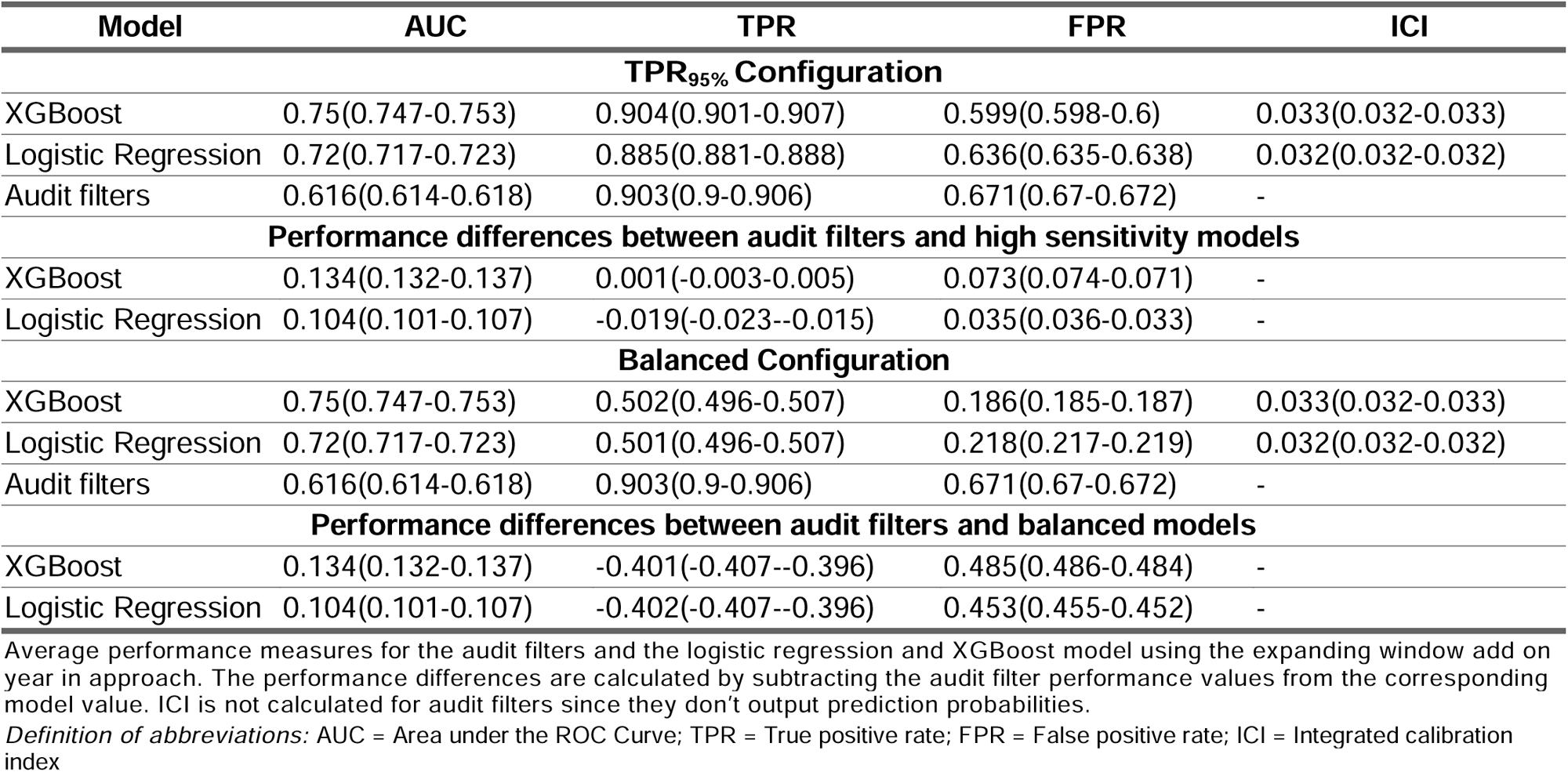
Performance Metrics.

### Model Development, Specification and Performance

The frequency of opportunities for improvement varied between 2017 and 2022, with the highest occurring in 2017 (n=112, 9%) and the lowest in 2018 (n=36, 3%). Annual characteristics are provided in eTable 4 (Supplement E2). Figure 2 provides the number of patients and opportunities for improvement for each year and corresponding training datasets.

**Figure 2.**
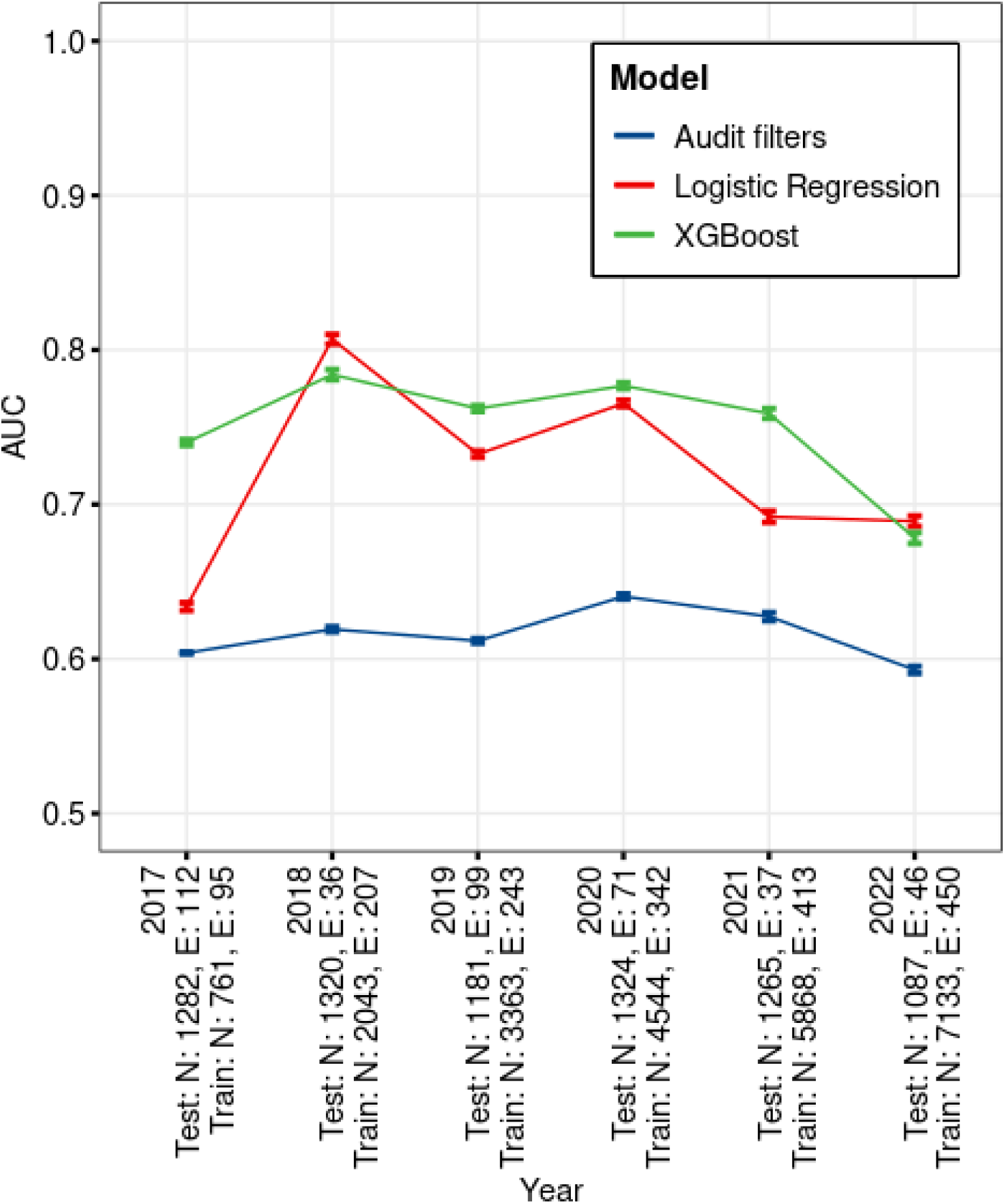
Annual AUC values for each model. Sample sizes and OFI number for each training and test set. Year wise model performance and sample sizes for the expanding window add one year in analysis. A) Mean area under the curve (AUC) per model and year. Lines represent 95% confidence intervals. For any given year, the AUCs were calculated with that year as the test set and all preceding years, starting with 2013, as the training set. For example, for 2019 the AUCs were calculated using 2019 as the test set and 2013-2018 as the training set. B) Opportunities for improvement (OFI) and sample sizes per year. The training sample and the test sample sizes includes the OFI in each sample respectively. For any given year, the training OFI and training sample size rows are the total number of patients with OFI and total number of patients in all preceding years respectively. The test OFI test sample size rows are the number of patients with OFI and the total number of patients in a specific year. For example, for 2019 the training OFI is the total number of patients with OFI 2013-2018, the training sample size is the total number of patients 2013-2018, the test OFI is the number of patients with OFI in 2019 and the test sample size the number of patients in 2019.

ISS was the most important predictor, followed by highest level of care. Figure 3 shows the average predictor importance for all years between 2017 and 2022 for all predictors.

**Figure 3.**
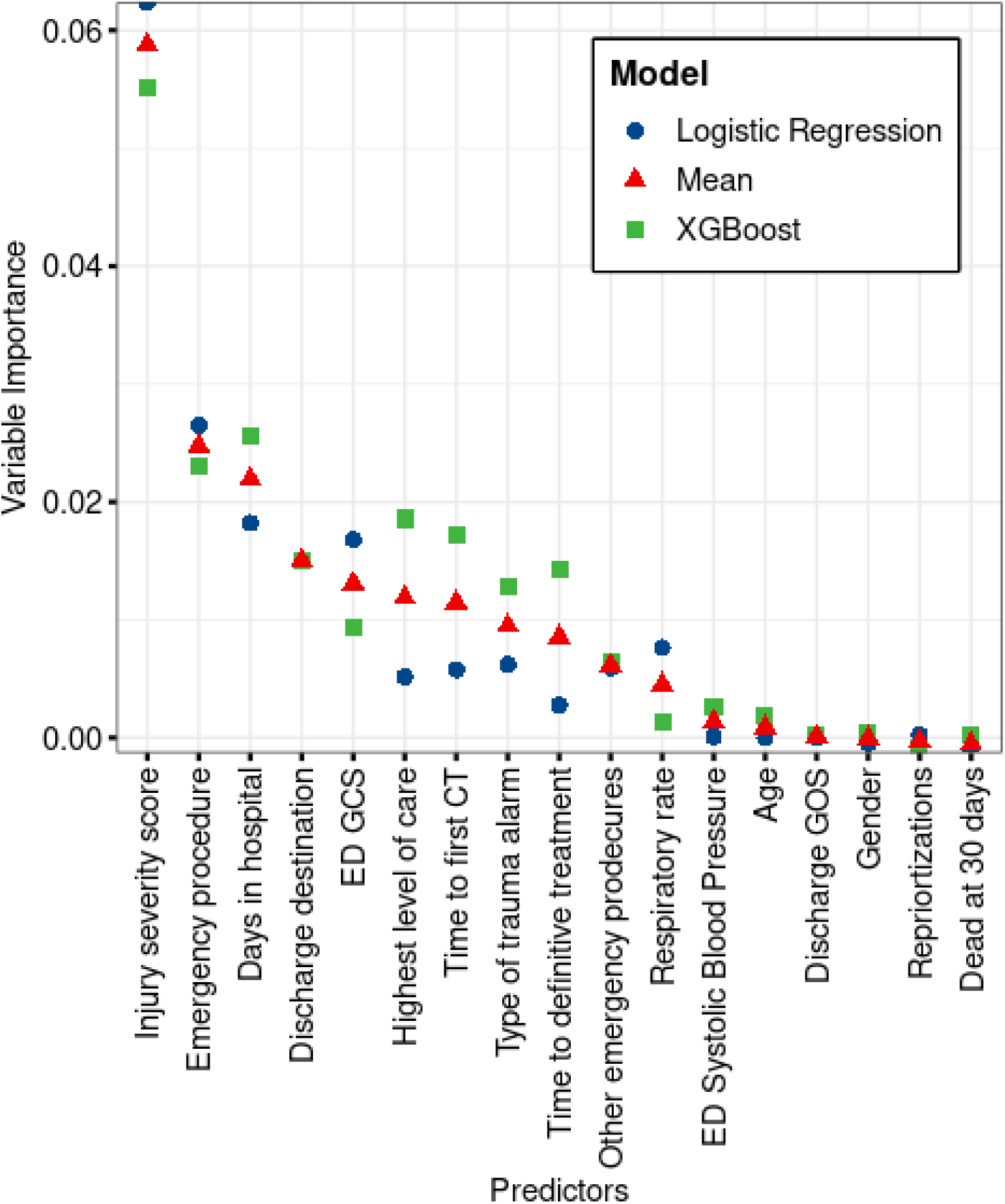
Average permuted variable importance for each model. The calculated, model-agnostic, permuted feature importance calculated using the AUC as the scoring metric. Variable importance is measured as AUC change when a variable is permuted. The model values are the average within that model between the years 2017 and 2022. The “Mean” value is the mean of all models. Definition of abbreviations: PH = pre-hospital; ED = emergency department, GCS = Glasgow Coma Scale, GOS = Glasgow Outcome Scale.

The logistic regression and XGBoost models were well calibrated with ICIs (95% CI) of 0.032 (0.032-0.032) and 0.033 (0.032-0.033). When averaging the results from each year, the audit filters had an AUC (95% CI) of 0.616 (0.614-0.618), a TPR (95% CI) of 0.903 (0.9-0.906), and a FPR (95% CI) of 0.671 (0.67-0.672). Compared to the audit filters, both the logistic regression and XGBoost models had higher AUCs (95% CI) of 0.72 (0.717-0.723) and 0.75 (0.747-0.753).

The XGBoost based model had a significantly lower FPR (95% CI) of 0.599 (0.598-0.6) while still retaining a superior TPR (95% CI) of 0.904 (0.901-0.907). The logistic regression model similarily displayed a superior FPR (95% CI) of 0.636 (0.635-0.638), however to the cost an inferior TPR (95% CI) of 0.885 (0.881-0.888).

In the TPR_95%_ configuration, the XGBoost model achieved a TPR (95% CI) of 0.904 (0.901-0.907) with a significantly lower FPR (95% CI) of 0.599 (0.598-0.6) compared to audit filters. The logistic regression model achieved a TPR (95% CI) of 0.885 (0.881-0.888) and a FPR (95% CI) of 0.636 (0.635-0.638). Both models demonstrated good calibration, with ICIs (95% CI) of 0.033 (0.032-0.033) for XGBoost and 0.032 (0.032-0.032) for logistic regression. In the balanced configuration, the XGBoost model had a TPR (95% CI) of 0.502 (0.496-0.507) and a FPR (95% CI) of 0.186 (0.185-0.187). The logistic regression model showed a TPR (95% CI) of 0.501 (0.496-0.507) and a FPR (95% CI) of 0.218 (0.217-0.219).

Figure 3 shows annual AUC values between 2017 and 2022. eFigures 1, eFigure 2 and eFigure 3 in the supplemental shows the annual TPR, FPR values and receiver operating characteristic curves between 2017 and 2022.

## Discussion

Both the XGBoost and logistic regression prediction models outperformed audit filters in predicting opportunities for improvement among adult trauma patients, with XGBoost showing the best overall performance. The performances of both models were modest, and the audit filters exhibited poor performance. Unlike audit filters, these models can be configured towards specific goals where we tested two configuration strategies: one prioritizing a higher TPR with a moderate reduction in FPR, and another accepting a moderate loss in TPR for a substantial reduction FPR. This adaptability allows these models to better balance identifying opportunities for improvement and managing the screening burden, which in combination with a superior overall performance offers potential advantages over traditional audit filters.

### Limitations

Importantly, our models’ results are most likely falsely low due to two limitations. First, the add on year in approach to simulate prospective implementation resulted in small sample sizes between 2017 and 2020, leading to poorer configuration. Second, these models are only evaluated against opportunities for improvement identified within the current peer review system. The low frequency of opportunities for improvement in this study, compared to previous studies, suggests potential false negatives (10,25,26). These false negatives would favor the models and increase their performance, as they were missed by the current audit filter and peer review system.

While defined as a binary variable, opportunities for improvement includes a diverse set of outcomes ranging from preventable deaths to lacking communication. The heterogeneity of these outcomes represents a range of clinical events, each likely correlating to different predictors. In addition, machine learning models struggle to handle rare events, and despite being an aggregate of all previously identified errors, the opportunities for improvement frequency is only 6%; as a result, opportunities for improvement is a considerable predictive challenge.

Another potential risk is a “data shift”. Due to feasibility, mortality and morbidity reviews and corresponding corrective actions can focus only on a subset of opportunities for improvement at any given time. Hence, a correctly flagged opportunities for improvement might not be registered since the system must prioritize other areas in need of correction. If human resources could be removed from basic screening tasks by reducing false positives, they could possibly be allocated toward more in-depth reviews, reducing the need to prioritize opportunities for improvement subgroups.

### Interpretation and generalisability

While the use of audit filters when screening for opportunities for improvement remain the current state-of-the-art technique for trauma quality improvement programs their effectiveness, especially in in the mature trauma system, have long been questioned (8,9). The static nature of audit filters makes them less effective as the trauma system adapts, potentially requiring frequent changes over time. The adaptive nature of machine learning models offers a promising solution, allowing the models and subsequently selected patient cases to change as the models develop over time. While our study suggests this as a possibility, prospective implementation is needed for true evaluation.

The XGBoost model in the TPR_95%_ configuration had a similar TPR to that of the audit filters but achieved a 11% (n=90) reduction in the annual screening burden. This reduction in false positive are further highlighted when configuring toward balanced performance, reducing the screening burden by 72% (n=572) while identifying 46% (n=28) fewer opportunities for improvement annually. Although the reduction in TPR is not ideal, this trade-off should be considered given that trauma systems may forgo peer review altogether due to the high FPR of audit filters. Thus, the significant reduction in FPR could offer benefits for settings with limited resources.

Additionally, the need for extra human review as a consequence of the high FPR before the mortality and morbidity review risks introducing bias, reducing the intended multidisciplinary approach. Mortality prediction models have been suggested as a solution; however they perform poorly and are only applicable in mortality-related cases (10). The balanced models offer a potential solution where over 80% of the flagged cases contain opportunities for improvement. Cases can therefore be brought directly from standard trauma registries to the mortality and morbidity review without additional human pre-screening. The possibility to configure these models therefore represent a tool for high-yield selection in context that want to include morbidity cases while protecting the intended multidisciplinary approach of the final review.

A systematic review and meta-analysis by Zhang et al. investigated the performance of different machine learning applications and learners in the trauma setting found that similar performance where often found using Logistic regression compared to more complex machine learning models (28). Our study showed that XGBoost had a small, but significant, performance advantage compared to logistic regression however both performed modestly. Instead, a substantial performance increase would probably require both higher quantity and quality of data, e.g., higher-resolution data such as vital sign series and defined, complete and consistent opportunities for improvement classifications. However, in doing so one sacrifices external validity and general feasibility compared to the models in our study, which are easily applicable in settings with standard registries following the Utstein template (14).

## Conclusion

It is important to note that perfect performance is far from expected. Comparing these models to entire systems using a combination of quantitative screening and several human reviews, including a multidisciplinary review, is unfair and not the goal of this paper. Instead, we strive to facilitate quality improvement efforts through a combination of human and artificial intelligence. Compared to audit filters, these models offer increased overall performance and the option to balance and optimize the tradeoff between screening burden and sensitivity goals, giving each trauma quality improvement program the potential to standardize and automate part of the review system in a way that complements human efforts.

## Data Availability

The data that support the findings of this study are available following the approval of a project suggesting to use the data by the Swedish Ethical Review Authority and the appropriate bodies at the Karolinska University Hospital. More information is available on request from the corresponding author, J. Attergrim.

## Acknowledgments

The authors thank Liselott Västerbo for her participation in collecting and recording the data and screening for opportunities for improvement. The authors also thank all professionals who participated in the monthly mortality and morbidity conferences.

## Contributors

M.G.W. and J.A. obtained funding and conceptualized the study. M.G.W., J.A. and K.S. drafted the study protocol. M.G.W., J.A., K.S. and M.J. wrote the statistical analysis plan. J.A. and K.S. performed the statistical analysis and model development. J.A. and K.S. drafted the manuscript, which was critically revised by all the authors. All the authors read and approved the final manuscript. J.A., K.S. and M.G.W. are guarantors. J.A. and K.S. contributed equally to this work. The corresponding author attests that all listed authors meet authorship criteria and that no others meeting the criteria have been omitted.

## Competing interests

All the authors have completed the ICMJE uniform disclosure form at http://www.icmje.org/disclosure-of-interest/ and declare the following: M.G.W. and J.A. received grants related to this study from the Swedish Society of Medicine and from “The Swedish Carnegie Hero Fund”. The authors have no financial relationships with any organizations that might have an interest in the submitted work in the previous three years. The authors have o other relationships or activities could appear to have influenced the submitted work.

The lead author (the manuscript’s guarantors) affirms that the manuscript is an honest, accurate, and transparent account of the study being reported; that no important aspects of the study have been omitted; and that any discrepancies from the study as planned (and, if relevant, registered) have been explained.

Dissemination to participants and related patient and public communities:

The results will be disseminated through local and international conferences. To date, results have presented at the London Trauma Conference (December 2022). Additionally, code for replicating the results and models are publicly available: https://github.com/noacs-io/predicting-ofi-in-trauma

