## Supplemental text, tables and figures for "Predicting Opportunities for Improvement in Trauma Care: A Registry-Based Cohort Study"

**Predicting Opportunities for Improvement in Trauma Care Using Machine Learning**

### Supplement E1. Learners and models

**Audit filter model**

The Karolinska University’s trauma care quality database includes binary results for each audit filter on a trauma patient case. For example, a nurse reviewing a patient might identify a reason for the time to computed tomography being more than 30 minutes from arrival to the hospital and thus flag that audit filter as negative. As a result, determining the true performance of the audit filters diﬀicult. To address this, a model was created that simulates the use of audit filters by creating new predictors for each audit filter using registry data. If at least one of these new predictors is positive, the audit filter model predicts the presence of an OFI with a 100% probability. However, the trauma registry does not include data for the last four audit filters in the list above; therefore, the results from the trauma care quality database, which includes some manual manipulation and data not available to the other models, were used in the model instead. This information was not used by the ML models.

**Logistic Regression**

Logistic regression (LR) is a statistical method used to model a binary dependent variable using independent variables, also known as “predictors”. These predictors can be continuous or categorical. In multivariable logistic regression, more than one predictor is used. The logistic function, which maps the input to a value between 0 and 1, is used to calculate the probability that a specific combination of predictor values corresponds to one of the binary values of the dependent variable. To find the coeﬀicients for the model, a linear equation is created by plotting the logistic function of the dependent variable against the predictor. The intercept is calculated at the point where the predictor is 0, and the slope is calculated using the log of the odds and the odds ratio for continuous and categorical predictors, respectively. Fitting is done by calculating the maximum likelihood. The T-square test is used to determine the significance of continuous variables, and the chi-square test or Fisher’s test is used for categorical predictors.

**K-Nearest Neighbor**

K-nearest neighbors (k-NN) is a nonparametric supervised machine learning method that

was originally developed by Evelyn Fix and Joseph Hodges in 1951 and later expanded upon by Thomas Cover in 1967 (31). It can be used for both classification and regression tasks. The method works by calculating the distances between a query and all examples in the data and selecting the k number of examples that are closest to the query. Classification is performed by choosing the most frequent label among the selected examples, while regression is performed by calculating the average of the labels of the selected examples.

**Support Vector Machine**

Support-vector machines (SVMs) were first introduced in 1995 (27). SVMs are supervised learners that are useful for classification tasks. Given a training dataset, the model creates a function, namely, a point, line, plane or hyperplane, depending on the number of predictors. The function divides the data into two categories, with or without the outcome. The width halfway between the categories is called the threshold. The distance from the chosen observations to the threshold is called the soft margin. Using cross-validation, the function is altered to limit overfitting and to minimize misclassification and observation on the “wrong” side of the function while still maximizing the soft margin. This function is called a support vector classifier. If the classifier cannot divide the data, the SVM temporarily adds dimensions to the data to better split the data into observations with or without the outcome. This is called a kernel trick; several methods exist, including radial kernels or polynomial kernels.

**Decision Trees**

Decision trees (DTs) are commonly used for classification (classification tree) or numerical prediction (regression tree) (26). In this method, a tree-like structure is created, where each branch represents a binary statement based on a predictor and a threshold that can be true or false. The tree begins with the predictor that most accurately predicts the dependent variable. Several methods can be used to quantify this accuracy, including entropy, information gain, or the Gini impurity. Calculating Gini impurity is the most common method and is calculated as 1 - p1ˆ2 - p2ˆ2, where p1 and p2 represent the ending probabilities in each outcome. The

Gini impurity is then adjusted or weighted in relation to the total number of cases in that “node”. The first statement, called the root node, is the predictor with the most accurate Gini impurity. The subsequent predictors are called nodes, and the final nodes containing the classification are called leaf nodes. A node becomes a leaf node once all predictors are used or if the adjusted Gini impurity increases. For binary categorical predictors, the Gini impurity can be calculated directly. If the predictor is continuous, the cutoff value or average between values that gives the lowest Gini impurity must be found. If the predictor is categorical and has multiple levels, the Gini impurity must be calculated for each category and for all possible combinations.

**Random Forests**

The random forest (RF) model is a method used for classification or regression that involves the use of multiple decision trees (25). To create the decision trees, new datasets are first created using a bootstrapping approach. Instead of considering all the candidate predictors and calculating the Gini impurity, a random number of variables is selected, and the best variable is used. This process is repeated, removing previously used variables until the decision tree is completed. This process is then repeated to create multiple different trees. Additionally, the selected number of variables is randomized to create even more trees. When using a random forest for prediction, the majority outcome of all the decision trees is taken as the model’s prediction. This process of using bootstrapped data and taking the aggregate is called bagging. To estimate the accuracy, the out-of-bag dataset, which includes data that were not included in each bootstrap, can be run through the random forest algorithm. This accuracy is also called the out-of-bag error.

**XGBoost**

XGBoost, short for eXtreme Gradient Boosting, is an optimized distributed gradient boosting library that was first introduced in 2016 (28). It uses algorithms under the gradient boosting framework, offering parallel tree boosting to solve various problems, including classification. Boosting is a learning technique that builds strong classifiers by assembling a series of several weak classifiers. Unlike bagging algorithms, which mainly control for variance, boosting controls both bias and variance to minimize overfitting and optimize eﬀiciency. XGBoost builds on the principles of adaptive boosting (AdaBoost), which was the first booster developed for binary classification. AdaBoost is adaptive, meaning that subsequent classifiers are influenced by previous misclassifications. Multiple iterations are used to create a strong single composite learner by iteratively adding weak learners using a weighting vector adjusted by the previous learners’ misclassifications. Gradient boosting is a generalization of AdaBoost that aims to minimize the loss function by adding weak learners using a gradient descent optimization algorithm. XGBoost builds on these principles to create a faster, optimized model for use by several data scientists and for various machine learning goals, including classification.

**LightGBM**

The LightGBM algorithm is a decision tree algorithm that was first introduced in 2016 and originally developed by Microsoft (29). While it has many similarities to XGBoost, including parallel training, regularization, sparse optimization, bagging, and early stopping of the algorithm, it also provides additional eﬀiciency and memory consumption advantages, earning the name “light gradient-boosting machine”. LightGBM grows the tree leafwise and chooses the leaf estimated to yield the largest loss reduction. Unlike XGBoost, which uses sorted- based decision trees, LightGBM uses a histogram-based decision tree learning algorithm. These features allow LightGBM to run faster than other machine learning algorithms while maintaining accuracy.

**CatBoost**

Originally developed by Yandex, CatBoost (30) is a gradient boosting approach that attempts to solve categorical features using a permutation-driven alternative. It offers several features, including oblivious trees or symmetric trees for fast execution and ordered boosting to combat overfitting.

### Supplement E2. Tables

**eTable 1 :** Current audit filters at KUH

**A specialized nurse checks if any of the following audit criteria are met:**

In hospital death or death within 30-days.

Systolic blood pressure less than 90

Glasgow coma scale less than 9 and not intubated.

An Injury severity score greater than 15, but the patient was not admitted to the intensive care unit.

An Injury severity score greater than 15 and no trauma team activation.

Time to first major intervention more than 60 minutes from arrival at the hospital

Time to computed tomography more than 30 minutes from arrival at the hospital.

No anticoagulant therapy administered within 72 hours after a traumatic brain injury.

The presence of cardiopulmonary resuscitation with thoracotomy

The presence of a liver or spleen injury

Massive transfusion, defined as 10 or more units of packed red blood cells within 24 hours.

| **eTable 2** : Complete information on used predictors for ML models. | | | |
| --- | --- | --- | --- |
|  | **OFI (N=496)** | **No OFI (N=7813)** | **Overall (N=8309)** |
| **Age** |  |  |  |
| Mean (SD) | 49 (22) | 45 (21) | 45 (21) |
| Median [Min, Max] | 49 [15, 97] | 42 [15, 100] | 43 [15, 100] |
| **Sex** |  |  |  |
| Female | 136 (27%) | 2418 (31%) | 2554 (31%) |
| Male | 360 (73%) | 5395 (69%) | 5755 (69%) |
| **Dead at 30 days** |  |  |  |
| Yes | 41 (8%) | 686 (9%) | 727 (9%) |
| No | 453 (91%) | 7118 (91%) | 7571 (91%) |
| Missing | 2 (<1%) | 9 (<1%) | 11 (<1%) |
| **ED GCS** |  |  |  |
| Mean (SD) | 14 (3) | 14 (2) | 14 (2) |
| Median [Min, Max] | 15 [3, 15] | 15 [3, 15] | 15 [3, 15] |
| Missing | 49 (10%) | 817 (10%) | 866 (10%) |
| **Discharge Glasgow outcome scale** |  |  |  |
| Mean (SD) | 3 (1) | 4 (1) | 4 (1) |
| Median [Min, Max] | 3 [1, 5] | 4 [1, 5] | 4 [1, 5] |
| Missing | 0 (0%) | 1 (<1%) | 1 (<1%) |
| **ED Systolic Blood Pressure** |  |  |  |
| Mean (SD) | 133 (34) | 133 (33) | 133 (33) |
| Median [Min, Max] | 135 [0, 237] | 135 [0, 285] | 135 [0, 285] |
| Missing | 0 (0%) | 13 (<1%) | 13 (<1%) |
| **Respiratory rate** |  |  |  |
| Mean (SD) | 19 (5) | 18 (5) | 18 (5) |
| Median [Min, Max] | 18 [0, 40] | 18 [0, 60] | 18 [0, 60] |
| Missing | 51 (10%) | 818 (10%) | 869 (10%) |
| **Injury severity score** |  |  |  |
| Mean (SD) | 19 (11) | 12 (13) | 12 (13) |
| Median [Min, Max] | 17 [0, 75] | 9 [0, 75] | 9 [0, 75] |
| Missing | 0 (0%) | 11 (<1%) | 11 (<1%) |
| **Emergency procedure** |  |  |  |
| Thoracotomy | 8 (2%) | 97 (1%) | 105 (1%) |
| Laparotomy | 28 (6%) | 217 (3%) | 245 (3%) |
| Pelvis Packing | 0 (0%) | 5 (<1%) | 5 (<1%) |
| Revascularization | 12 (2%) | 37 (<1%) | 49 (1%) |
| Radiological intervention | 32 (6%) | 69 (1%) | 101 (1%) |
| Craniotomy | 42 (8%) | 242 (3%) | 284 (3%) |
| Intracranial pressure measurement | 13 (3%) | 90 (1%) | 103 (1%) |
| Other | 131 (26%) | 1313 (17%) | 1444 (17%) |
| Missing | 230 (46%) | 5743 (74%) | 5973 (72%) |
| **Other emergency procedures** |  |  |  |
| Chest drain | 40 (8%) | 400 (5%) | 440 (5%) |
| External fixation of fracture | 22 (4%) | 165 (2%) | 187 (2%) |
| Major fracture surgery | 37 (7%) | 302 (4%) | 339 (4%) |
| Wound revision in OR | 27 (5%) | 391 (5%) | 418 (5%) |
| Other action | 5 (1%) | 56 (1%) | 61 (1%) |
| Missing | 365 (74%) | 6499 (83%) | 6864 (83%) |
| **Time to first CT** |  |  |  |
| Mean (SD) | 76 (129) | 71 (159) | 71 (157) |
| Median [Min, Max] | 40 [6, 1339] | 33 [0, 7073] | 33 [0, 7073] |
| Missing | 42 (8%) | 956 (12%) | 998 (12%) |
| **Time to first major intervention** |  |  |  |
| Mean (SD) | 271 (323) | 251 (353) | 253 (349) |
| Median [Min, Max] | 143 [3, 1420] | 101 [0, 2036] | 106 [0, 2036] |
| Missing | 230 (46%) | 5743 (74%) | 5973 (72%) |
| **Highest level of care** |  |  |  |
| Emergency department | 22 (4%) | 1478 (19%) | 1500 (18%) |
| General ward | 116 (23%) | 2962 (38%) | 3078 (37%) |
| Operation Theatre | 141 (28%) | 1454 (19%) | 1595 (19%) |
| Specialist ward/Intermediate ward | 50 (10%) | 343 (4%) | 393 (5%) |
| Intensive care unit | 167 (34%) | 1576 (20%) | 1743 (21%) |
| **Type of trauma alarm** |  |  |  |
| Trauma alarm level 1 | 196 (40%) | 4133 (53%) | 4329 (52%) |
| Trauma alarm level 2 | 81 (16%) | 1907 (24%) | 1988 (24%) |
| No trauma alarm | 53 (11%) | 551 (7%) | 604 (7%) |
| Missing | 166 (33%) | 1222 (16%) | 1388 (17%) |
| **Reprioritizations** |  |  |  |
| no reprioritization | 313 (63%) | 6387 (82%) | 6700 (81%) |
| reprioritization to level 1 | 15 (3%) | 152 (2%) | 167 (2%) |
| reprioritization to level 2 | 2 (<1%) | 43 (1%) | 45 (1%) |
| Alarm cancelled | 0 (0%) | 9 (<1%) | 9 (<1%) |
| Missing | 166 (33%) | 1222 (16%) | 1388 (17%) |
| **Days in hospital** |  |  |  |
| Mean (SD) | 12 (14) | 6 (14) | 7 (14) |
| Median [Min, Max] | 7 [1, 109] | 3 [1, 392] | 3 [1, 392] |
| **Discharge destination** |  |  |  |
| Home | 199 (40%) | 4902 (63%) | 5101 (61%) |
| Rehab | 199 (40%) | 1394 (18%) | 1593 (19%) |
| Morgue | 35 (7%) | 614 (8%) | 649 (8%) |
| ICU (higher care level) | 2 (<1%) | 42 (1%) | 44 (1%) |
| ICU (same care level) | 10 (2%) | 113 (1%) | 123 (1%) |
| Other department | 38 (8%) | 440 (6%) | 478 (6%) |
| psychiatric care | 13 (3%) | 303 (4%) | 316 (4%) |
| Missing | 0 (0%) | 5 (<1%) | 5 (<1%) |
| Definition of abbreviations: OFI = Opportunity for Improvement; ED = Emergency Department; GCS = Glasgow Coma Scale; PH = Pre Hospital; Time to first CT and Time to first major intervention : Measured in minutes from arrival at the hospital | | | |

| **eTable 3:** Number of specific Opportunity for improvement categories | |
| --- | --- |
| **Category (n)***^1^* | **Opportunities For Improvement, N = 496** |
| **Preventable death (42)**  Mortality that could have been prevented in the optimal trauma care system under optimal treatment conditions as decided by the MoM*^2^* -review. | **Preventable death**: 4  **Possible preventable death:** 38 |
| **Missed diagnosis (68)**  Failure to appropriately diagnose an injury or condition despite having sufficient information and resources. | **Missed injury**: 67  **Tertiary survey**: 1 |
| **Delay in treatment (72)**  Inappropriate delays from arrival to treatment or imaging leading to harm. | **Delay to surgery**: 47  **Delay to CT***^3^*: 25 |
| **Clinical judgment error (176)**  An incorrect management strategy, despite sufficient information and resources, leading to patient harm. | **Triage in the ED***^2^*: 64  **Level of care**: 48  **Patient management**: 46  **Communication**: 16 |
| **Inadequate protocols (24)**  Lacking or not following protocols/guidelines resulting in patient harm. | **Trauma criteria/guidelines**: 22  **Inadequate routine**: 2 |
| **Inadequate resources (111)**  Lack of resources, including personnel, or material, leading to patient harm. | **Competence**: 2  **Resources**: 73  **Logistics/technical**: 36 |
| **Other errors (3)** | **Other**: 1  **Patient management/logistics**: 1  **Prehospital management**: 1 |
| *^1^* Definitions of categories of Opportunities for Improvements (OFI) used. *^2^* MoM = Mortality and morbidity,  *^3^*CT = Computed tomography, ED = Emergency Department | |

| **eTable 4.** Demographic and Clinical Characteristics of patients screened for OFI.  37 | | | | | | | | |
| --- | --- | --- | --- | --- | --- | --- | --- | --- |
|  | **2013-2016 (N=761)** | **2017 (N=1282)** | **2018 (N=1320)** | **2019 (N=1181)** | **2020 (N=1324)** | **2021 (N=1265)** | **2022 (N=1087)** | **Overall (N=8220)** |
| **OFI** |  |  |  |  |  |  |  |  |
| Yes | 95 (12%) | 112 (9%) | 36 (3%) | 99 (8%) | 71 (5%) | 37 (3%) | 46 (4%) | 496 (6%) |
| No | 666 (88%) | 1170 (91%) | 1284 (97%) | 1082 (92%) | 1253 (95%) | 1228 (97%) | 1041 (96%) | 7724 (94%) |
| **Age** |  |  |  |  |  |  |  |  |
| Mean (SD) | 50 (23) | 45 (21) | 44 (20) | 45 (21) | 44 (21) | 45 (21) | 47 (22) | 45 (21) |
| Median [Min, Max] | 48 [15, 99] | 42 [15, 98] | 41 [15, 98] | 43 [15, 97] | 41 [15, 100] | 43 [15, 100] | 45 [15, 97] | 43 [15, 100] |
| **Sex** |  |  |  |  |  |  |  |  |
| Female | 225 (30%) | 408 (32%) | 418 (32%) | 348 (29%) | 384 (29%) | 389 (31%) | 352 (32%) | 2524 (31%) |
| Male | 536 (70%) | 874 (68%) | 902 (68%) | 833 (71%) | 940 (71%) | 876 (69%) | 735 (68%) | 5696 (69%) |
| **Dead at 30 days** |  |  |  |  |  |  |  |  |
| Yes | 232 (30%) | 92 (7%) | 73 (6%) | 88 (7%) | 83 (6%) | 74 (6%) | 76 (7%) | 718 (9%) |
| No | 527 (69%) | 1184 (92%) | 1247 (94%) | 1092 (92%) | 1241 (94%) | 1189 (94%) | 1011 (93%) | 7491 (91%) |
| Missing | 2 (<1%) | 6 (<1%) | 0 (0%) | 1 (<1%) | 0 (0%) | 2 (<1%) | 0 (0%) | 11 (<1%) |
| **Highest level of care** |  |  |  |  |  |  |  |  |
| Emergency department | 37 (5%) | 316 (25%) | 357 (27%) | 232 (20%) | 212 (16%) | 176 (14%) | 159 (15%) | 1489 (18%) |
| General ward | 185 (24%) | 467 (36%) | 469 (36%) | 413 (35%) | 528 (40%) | 567 (45%) | 407 (37%) | 3036 (37%) |
| Operation Theatre | 196 (26%) | 227 (18%) | 237 (18%) | 222 (19%) | 263 (20%) | 218 (17%) | 216 (20%) | 1579 (19%) |
| Specialist ward/Intermediate ward | 23 (3%) | 21 (2%) | 37 (3%) | 61 (5%) | 78 (6%) | 82 (6%) | 84 (8%) | 386 (5%) |
| Intensive care unit | 320 (42%) | 251 (20%) | 220 (17%) | 253 (21%) | 243 (18%) | 222 (18%) | 221 (20%) | 1730 (21%) |
| **Injury severity score** |  |  |  |  |  |  |  |  |
| Mean (SD) | 24 (17) | 11 (12) | 10 (12) | 11 (12) | 11 (12) | 11 (12) | 12 (12) | 12 (13) |
| Median [Min, Max] | 21 [1, 75] | 9 [0, 75] | 5 [0, 75] | 9 [0, 75] | 9 [0, 75] | 9 [0, 75] | 9 [0, 75] | 9 [0, 75] |
| Missing | 0 (0%) | 3 (<1%) | 0 (0%) | 1 (<1%) | 3 (<1%) | 1 (<1%) | 2 (<1%) | 10 (<1%) |
| **Time to first CT** |  |  |  |  |  |  |  |  |
| Mean (SD) | 82 (304) | 86 (129) | 71 (119) | 66 (126) | 77 (172) | 58 (131) | 61 (123) | 71 (157) |
| Median [Min, Max] | 35 [10, 7073] | 49 [14, 1401] | 37 [0, 1403] | 32 [0, 1410] | 27 [0, 1428] | 27 [3, 1428] | 28 [4, 1380] | 33 [0, 7073] |
| Missing | 124 (16%) | 143 (11%) | 141 (11%) | 137 (12%) | 162 (12%) | 141 (11%) | 139 (13%) | 987 (12%) |
| **Time to** **first major intervention** |  |  |  |  |  |  |  |  |
| Mean (SD) | 219 (318) | 212 (306) | 236 (321) | 202 (297) | 282 (381) | 323 (413) | 299 (372) | 253 (349) |
| Median [Min, Max] | 108 [1, 1935] | 100 [0, 2036] | 110 [0, 1425] | 99 [0, 1422] | 105 [0, 1428] | 116 [2, 1432] | 110 [1, 1426] | 106 [0, 2036] |
| Missing | 333 (44%) | 1001 (78%) | 1027 (78%) | 864 (73%) | 972 (73%) | 929 (73%) | 772 (71%) | 5898 (72%) |
| OFI = Opportunity for Improvement; Time to first CT and Time to first major intervention : Measured in minutes from arrival at the hospital. | | | | | | | | |

| **eTable 5.** Complete average performance metrics for all tested ML models in the AOYI-analysis | | | | | | |
| --- | --- | --- | --- | --- | --- | --- |
| **Model** | **AUC** | **TPR** | **FPR** | **ICI** | **TP** | **FP** |
| **TPR_95%_ Configuration** | | | | | | |
| XGB | 0.75(0.747-0.753) | 0.904(0.901-0.907) | 0.599(0.598-0.6) | 0.033(0.032-0.033) | 60(60-60) | 700(701-698) |
| RF | 0.733(0.73-0.736) | 0.888(0.884-0.891) | 0.617(0.616-0.619) | 0.031(0.031-0.032) | 59(59-59) | 723(724-721) |
| CAT | 0.721(0.718-0.724) | 0.874(0.871-0.878) | 0.615(0.613-0.616) | 0.029(0.028-0.029) | 58(58-59) | 719(720-718) |
| LR | 0.72(0.717-0.723) | 0.885(0.881-0.888) | 0.636(0.635-0.638) | 0.032(0.032-0.032) | 58(57-58) | 745(746-743) |
| SVM | 0.715(0.712-0.717) | 0.863(0.86-0.867) | 0.592(0.591-0.593) | 0.035(0.035-0.036) | 58(58-58) | 691(692-690) |
| LGB | 0.714(0.711-0.717) | 0.89(0.887-0.893) | 0.682(0.681-0.683) | 0.031(0.03-0.031) | 60(59-60) | 796(797-795) |
| KNN | 0.697(0.694-0.7) | 0.878(0.874-0.881) | 0.675(0.674-0.677) | 0.033(0.033-0.034) | 59(59-59) | 791(792-789) |
| DT | 0.688(0.686-0.691) | 0.849(0.846-0.853) | 0.651(0.65-0.652) | 0.034(0.034-0.035) | 58(57-58) | 762(763-761) |
| Audit filter | 0.616(0.614-0.618) | 0.903(0.9-0.906) | 0.671(0.67-0.672) | - | 61(61-61) | 789(791-788) |
| **Performance differences between audit filters and high sensitivity models** | | | | | | |
| XGB | 0.134(0.132-0.137) | 0.001(-0.003-0.005) | 0.073(0.074-0.071) | - | -1(-1--1) | 90(88-91) |
| RF | 0.117(0.115-0.12) | -0.016(-0.02--0.011) | 0.054(0.055-0.052) | - | -2(-2--2) | 67(65-68) |
| CAT | 0.105(0.102-0.108) | -0.029(-0.033--0.024) | 0.057(0.058-0.055) | - | -3(-3--2) | 70(69-72) |
| LR | 0.104(0.101-0.107) | -0.019(-0.023--0.015) | 0.035(0.036-0.033) | - | -3(-4--3) | 45(43-46) |
| SVM | 0.099(0.096-0.101) | -0.04(-0.044--0.036) | 0.079(0.081-0.078) | - | -3(-3--3) | 98(97-100) |
| LGB | 0.098(0.095-0.101) | -0.013(-0.017--0.009) | -0.011(-0.01--0.012) | - | -1(-2--1) | -7(-8--5) |
| KNN | 0.081(0.078-0.084) | -0.025(-0.029--0.022) | -0.004(-0.003--0.005) | - | -2(-2--2) | -1(-3-0) |
| DT | 0.073(0.07-0.076) | -0.054(-0.058--0.049) | 0.02(0.022-0.019) | - | -3(-4--3) | 27(26-29) |
| **Balanced Configuration** | | | | | | |
| XGB | 0.75(0.747-0.753) | 0.502(0.496-0.507) | 0.186(0.185-0.187) | 0.033(0.032-0.033) | 33(33-33) | 218(219-217) |
| RF | 0.733(0.73-0.736) | 0.519(0.514-0.524) | 0.222(0.221-0.223) | 0.031(0.031-0.032) | 34(33-34) | 259(260-258) |
| CAT | 0.721(0.718-0.724) | 0.401(0.396-0.407) | 0.166(0.165-0.167) | 0.029(0.028-0.029) | 29(28-29) | 193(194-192) |
| LR | 0.72(0.717-0.723) | 0.501(0.496-0.507) | 0.218(0.217-0.219) | 0.032(0.032-0.032) | 34(33-34) | 257(258-256) |
| SVM | 0.715(0.712-0.717) | 0.522(0.516-0.527) | 0.239(0.238-0.24) | 0.035(0.035-0.036) | 35(35-36) | 282(283-281) |
| LGB | 0.714(0.711-0.717) | 0.52(0.514-0.525) | 0.241(0.24-0.242) | 0.031(0.03-0.031) | 35(35-35) | 281(282-280) |
| KNN | 0.697(0.694-0.7) | 0.497(0.492-0.502) | 0.245(0.244-0.246) | 0.033(0.033-0.034) | 35(35-36) | 285(286-283) |
| DT | 0.688(0.686-0.691) | 0.46(0.455-0.465) | 0.23(0.229-0.231) | 0.034(0.034-0.035) | 32(32-32) | 265(267-264) |
| Audit filter | 0.616(0.614-0.618) | 0.903(0.9-0.906) | 0.671(0.67-0.672) | - | 61(61-61) | 789(791-788) |
| **Performance differences between audit filters and balanced models** | | | | | | |
| XGB | 0.134(0.132-0.137) | -0.401(-0.407--0.396) | 0.485(0.486-0.484) | - | -28(-28--28) | 572(570-573) |
| RF | 0.117(0.115-0.12) | -0.384(-0.39--0.379) | 0.449(0.451-0.448) | - | -27(-28--27) | 531(529-532) |
| CAT | 0.105(0.102-0.108) | -0.502(-0.507--0.496) | 0.505(0.507-0.504) | - | -32(-33--32) | 597(595-598) |
| LR | 0.104(0.101-0.107) | -0.402(-0.407--0.396) | 0.453(0.455-0.452) | - | -27(-28--27) | 533(531-534) |
| SVM | 0.099(0.096-0.101) | -0.381(-0.387--0.376) | 0.433(0.434-0.432) | - | -26(-26--25) | 507(506-509) |
| LGB | 0.098(0.095-0.101) | -0.383(-0.389--0.378) | 0.431(0.432-0.429) | - | -26(-26--26) | 508(507-510) |
| KNN | 0.081(0.078-0.084) | -0.406(-0.412--0.401) | 0.426(0.428-0.425) | - | -26(-26--25) | 505(503-506) |
| DT | 0.073(0.07-0.076) | -0.443(-0.449--0.437) | 0.441(0.442-0.44) | - | -29(-29--29) | 524(522-525) |
| Average performance measures for all models and audit filters for the expanding window add on year in approach. The performance differences are calculated by subtracting the audit filter performance values from the corresponding model values. ICI is not calculated for audit filters since they don’t output prediction probabilities.  *Definition of abbreviations:* AUC = Area under the ROC Curve; FPR = False positive rate; ICI = Integrated calibration index; TP = True positive patients; FP = False positive patients; XGB = extreme gradiant booster; RF = Random Forest; CAT = CatBoost; LR = Logistic regression; SVM = Support Vector Machine; LGB = Light gradient booster; KNN = K-nearest neighbor; DT = Decision tree. | | | | | | |

### Supplement E3. Figures

ROC Curves for years 2017-2022

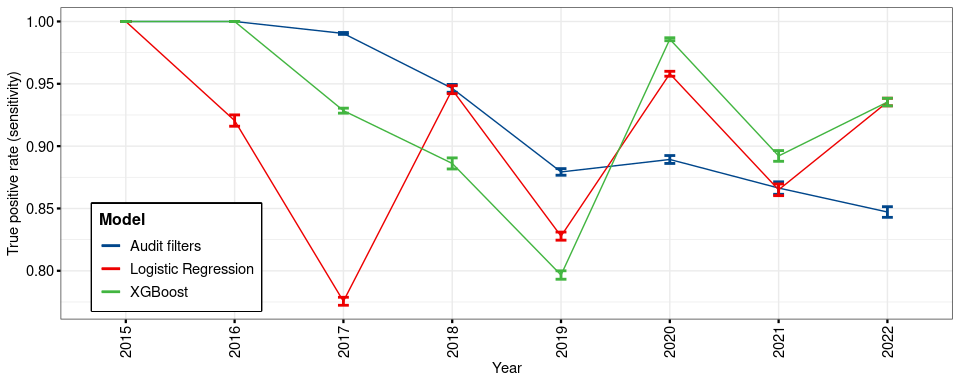

**eFigure 1**: Annual true positive rates

Legend: True positive rate values for each model and year

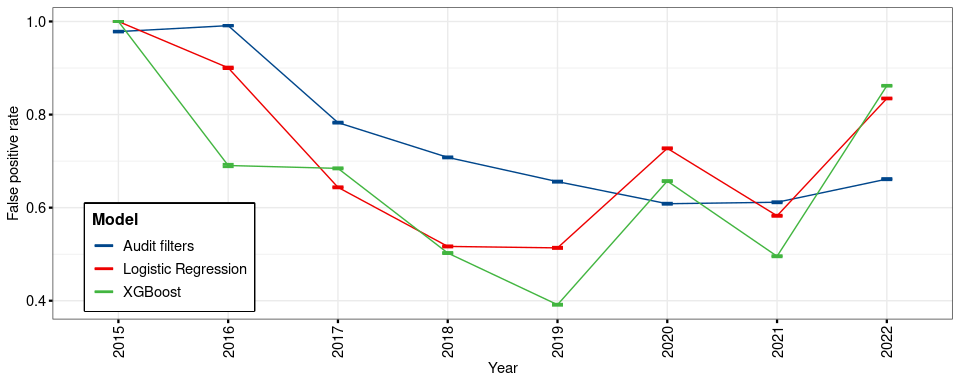

**eFigure 2:** Annual false positive rates

Legend: False positive rate values for each model and year

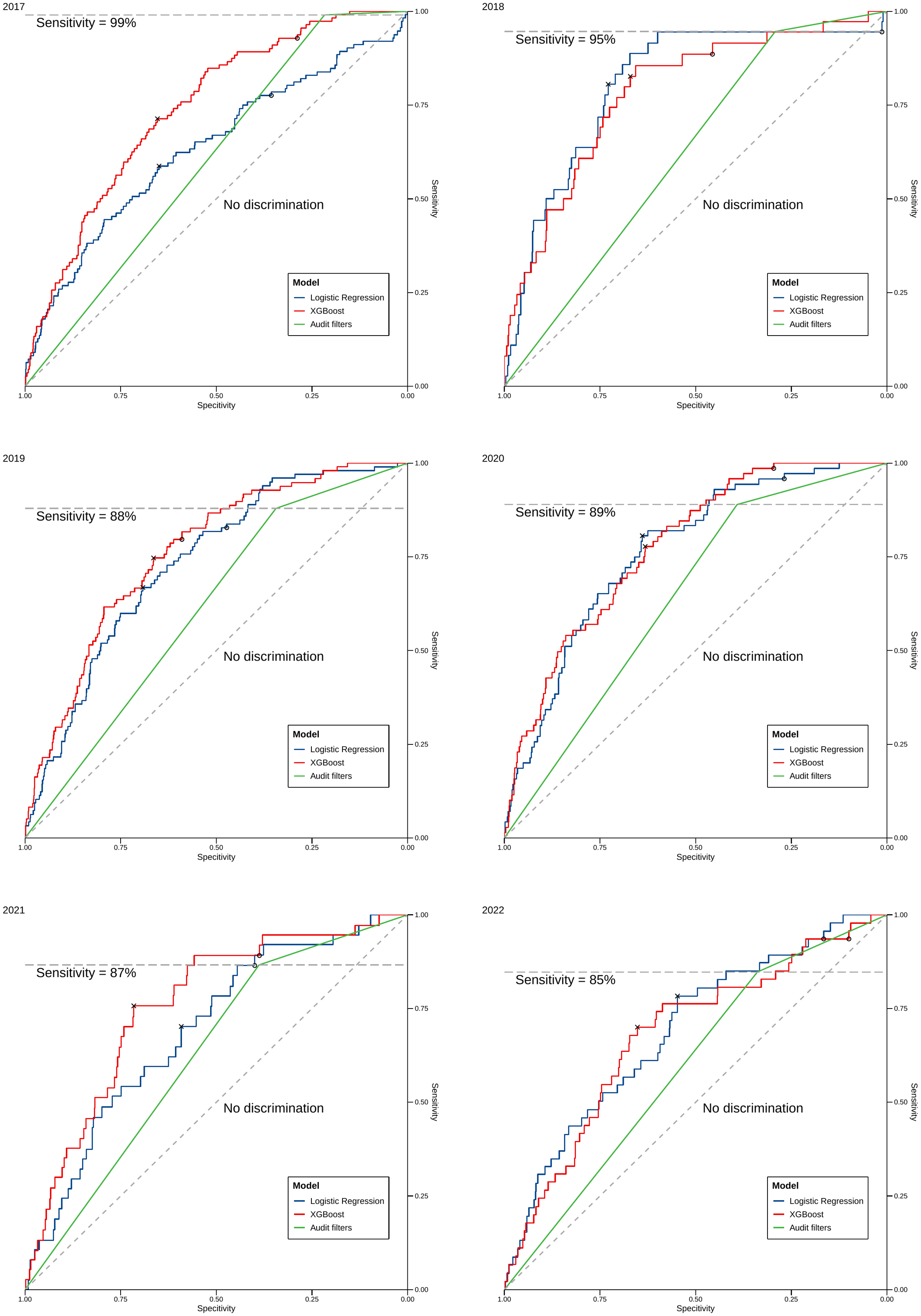

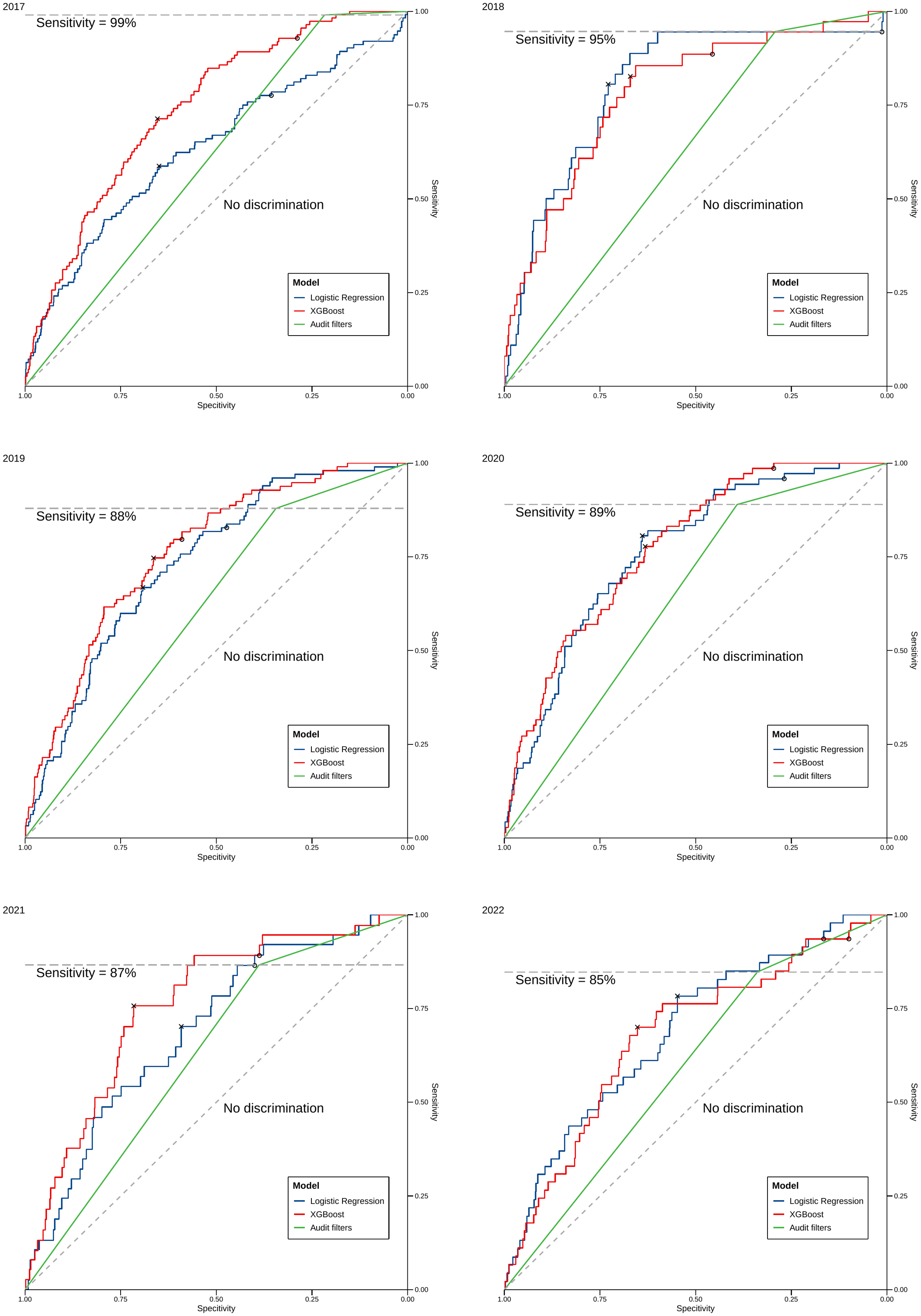

ROC Curves for years 2017-2022

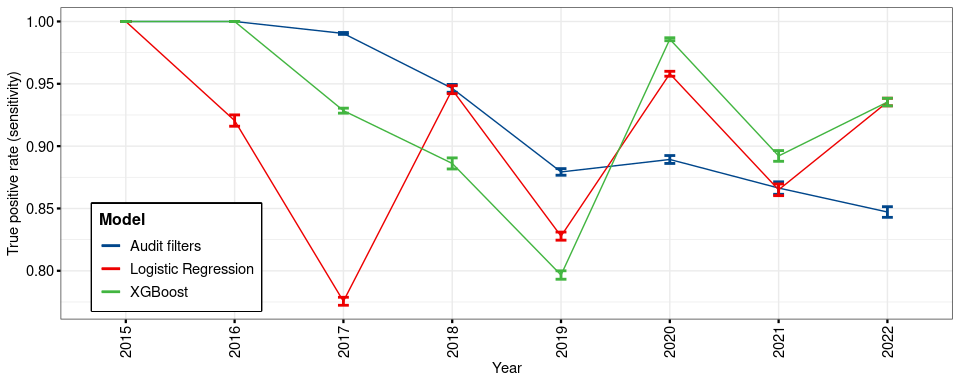

True positive rate values for each model and year

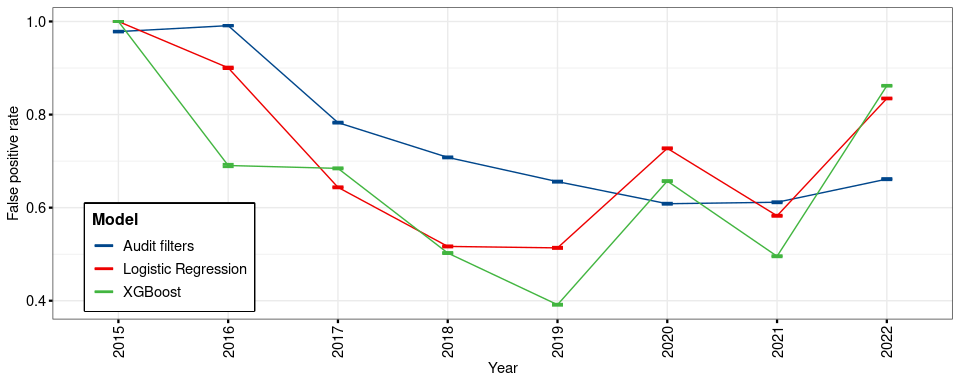

False positive rate values for each model and year

### Supplement E4. Figure Legends

**eFigure 1**. Annual specificity values for each model in the add on year in analysis.

**eFigure 2**. Annual sensitivity values for each model in the add on year in analysis.

**eFigure 3**. Receiver operating characteristic curves for all models when predicting OFI in the add on year in analysis.

**Legend:** OFI = Opportunity for improvement.
